# Generation of Risk Score for Serious Non-Steroidal Anti-Inflammatory Drug (NSAID) Induced Cardiovascular Events (NAÏVE) among Active-Duty Service Members and Veterans

**DOI:** 10.1101/2024.08.26.24312616

**Authors:** Timothy J Atkinson, Justin Petway, Whitney Forbes, Hannah Thorfinnson, Major Ryan C Costantino, Laura E Gressler

## Abstract

**Importance:** Cardiovascular disease (CVD) is a major global health concern, responsible for a significant proportion of deaths annually. Despite the widespread use of nonsteroidal anti-inflammatory drugs (NSAIDs) in managing various conditions, their association with cardiovascular events poses a challenge, and existing guidelines lack a reliable risk prediction tool. This study addresses the critical need for an evidence-based instrument to assess the likelihood of NSAID-induced cardiovascular events, that provides clinicians with valuable decision support to improve safety in their use for pain management, especially in patients vulnerable to cardiovascular events.

**Objective:** To develop a practical risk scoring tool, NSAID Induced Cardiovascular Events (NAÏVE), for estimating the risk of serious cardiovascular events associated with NSAID use.

**Design:** Retrospective nested case-control study

**Setting:** The study leveraged data from the DAVINCI database, integrating electronic health records, administrative data, and clinical data from both the Veterans Health Administration (VHA) and the Department of Defense (DoD).

**Participants:** The study cohort consisted of individuals with at least one NSAID pharmacy claim, with cases defined as those experiencing non-fatal myocardial infarction, non-fatal stroke, or new heart failure.

**Interventions:** Development of the NAÏVE risk scoring tool involved a comprehensive analysis of demographic, clinical, and prescription-related variables, including NSAID exposure, comorbidities, and medication history.

**Main Outcomes/Measures:** The primary outcome was the first occurrence of a cardiovascular event.

**Results:** The study cohort comprised 231,967 cases and 2,319,670 controls, identified from individuals with at least one NSAID pharmacy claim between October 1, 2016, and September 30, 2020. The risk index, NAÏVE, demonstrated strong discriminatory ability and calibration, with a C-statistic of 0.88. Variables such as age, NSAID exposure, comorbidities, and medication history were associated with increased odds of NSAID-induced cardiovascular events.

**Conclusions/Relevance:** NAÏVE is the first evidence-based risk scoring tool providing clinicians with valuable decision support for assessing the potential risk of serious cardiovascular events associated with NSAID use. It fills a crucial gap in clinical practice, allowing for transparent discussions with patients and shared decision-making regarding NSAID prescriptions. Further validation and prospective testing are warranted for broader applicability.

## Introduction

Cardiovascular disease (CVD) is a leading cause of morbidity and mortality worldwide, accounting for 31% of all global deaths or roughly 17.9 million deaths annually. CVD encompasses conditions such as myocardial infarction, stroke, and heart failure.^1^ Numerous studies have highlighted the prevalence of CVD and its associated risk factors which include hypertension, dyslipidemia, smoking, obesity, diabetes mellitus, sedentary lifestyle, and family history of CVD.^2^

Nearly 20 years ago, nonsteroidal anti-inflammatory drugs (NSAIDs) emerged as a threat to provoke cardiovascular events, and yet we still cannot accurately predict the risk of cardiovascular events associated with initiation or continuation of NSAID pharmacotherapy.^3-9^ NSAID-related cardiovascular risk has been evaluated in numerous studies with conflicting results mostly focusing on NSAID-specific factors such as COX selectivity, dose-dependent effects, and duration of NSAID therapy.^5,8,10-13^ Unlike other NSAID-related adverse effects such as renal function or GI ulceration/bleeding, previous studies determined that the cardiovascular risks associated with NSAID use do not appear related to dose or duration of treatment.^13,14^

Current clinical guidance on NSAID utilization revolves around avoidance of NSAIDs after a cardiovascular event and in certain disease states susceptible to NSAID-related adverse effects.^15-18^ However, for more than a hundred years, NSAIDs have been first-line medications for every arthritic and inflammatory condition making them one of the most utilized classes of medications worldwide and a cornerstone of treatment.^19^ Today, multiple evidence-based practice guidelines continue to recommend NSAIDs as first-line treatment making avoidance of NSAIDs impractical for many patients.^20-29^ NSAIDs are also important non-opioid pharmacotherapy options and key alternatives to opioid medications for many chronic pain conditions amid the opioid overdose crisis.^30,31^

Predictive models and scoring systems (risk indices) that estimate level of risk of an adverse outcome are commonly developed in medical research and clinical practice with the goal of preventing or mitigating an outcome. Common examples include cardiovascular disease^2^ and opioid-induced respiratory depression or overdose.^32^ While there are screening instruments to assess risk of cardiovascular disease in general, no published instruments currently provide clinically useful, evidence-based risk information about the likelihood of NSAID-induced cardiovascular events.

This study is a collaboration between clinicians from the Veterans Health Administration (VHA) and Enterprise Intelligence and Data Solutions Program Office of the Defense Healthcare Management Systems in the Department of Defense (DoD). We previously examined the potential predictors of serious NSAID-induced cardiovascular events in a case-control study of US active-duty military and Veterans. Factors with the strongest positive associations included age, NSAID-exposure, previous cardiovascular events, aspirin or other anticoagulants, and specific comorbidities.^33^ Based on the results from the previous study, a practical risk scoring tool was developed to estimate the likelihood of nonsteroidal anti-inflammatory drug-induced cardiovascular events (NAÏVE).

## Methods

### Study Design and Data Source

A retrospective nested case-control of the DAVINCI (Data Analysis and Visualization Initiative) data was leveraged to develop the risk scores for cardiovascular events.

DAVINCI is a collaborative effort between the Department of Defense (DoD) and the Veterans Health Administration (VHA) within the Department of Veterans Affairs (VA) in the US.^34^ The DAVINCI database integrates and analyzes various types of health data, including electronic health records (EHRs), administrative data, and clinical data from both the DoD and the VA using Observational Medical Outcomes Partnership (OMOP) common data models. The clinical records include information regarding healthcare visits, conditions, dispensed drugs, and procedures in both the inpatient and outpatient settings.

### Study Cohort

The study utilized the same derived cohort from the DAVINCI database as a previous study that identified factors associated with the cardiovascular events.^33^ More specifically, the study cohort consisted of individuals with at least one NSAID pharmacy claim with a days’ supply greater than 7 days between October 1, 2016 and September 30, 2020 and excluded individuals under the age of 18, with missing sex, or missing race variables. The authors determined that a 7 days’ supply requirement balances competing priorities to remove short-term prescriptions while recognizing that previous studies indicated that CV risk may be significant within the first 30 days. A total of 4,408,315 individuals were identified in the dataset with at least one NSAID pharmacy claim. Of these, 231,967 individuals were identified as cases. Cases were defined as individuals who experienced a cardiovascular event. A cardiovascular event included a non-fatal myocardial infarction, non-fatal stroke, and new onset of heart failure and was defined using International Classification of Disease, 10^th^ Revision, Clinical Modification (ICD10-CM) codes. Ten control patients were randomly assigned to each case and assigned the index date of the assigned case.^35,36^ To ensure that included individuals were regular users of TRICARE or VHA health services, individuals were required to have a recorded encounter in the 180 to 365 days prior to their identified or assigned index date. The study cohort consisted of 231,967 cases and 2,319,670 controls.

### Covariates and Outcomes

The first occurrence of a cardiovascular event, as defined by ICD-10 codes was the primary outcome of interest. Independent variables considered for the risk score were demographic, clinical, and prescription-related variables. Demographic variables were age, sex, and race. Comorbidity measures included diabetes^2^, hypertension^2^, dyslipidemia^2^, history of myocardial infarctions^1^, arthritis or spondylitis^37^, peripheral artery disease^2^, chronic kidney disease^1^, atherothrombotic disease^1^, history of tobacco use^2^, cerebrovascular disease^2^, coronary artery disease^2^, cardiomyopathy^1^, obstructive sleep apnea (OSA)^1^, liver dysfunction^1^, and chronic obstructive pulmonary disease (COPD).^1^ Prescription use measures included the use of aspirin and other anticoagulants. NSAID-specific drug information included NSAID selectivity (Cox 1, Cox 2, and non-selective),^5,10,14,38^ dosage (low/medium and high dose),^10,13,14,39^ and time since initial exposure (≤30, 31-90, 91+ and no exposure).^6,13,14,39^ Independent variables were collected in the 180 days prior to the index date.

### Statistical Analyses

The sample was characterized using descriptive statistics. Continuous variables were summarized using means and interquartile ranges and compared between cases and controls using t-tests. Categorical variables were summarized using frequencies and proportions. Categorical variables between cases and controls were compared using Chi-square tests.

Similar to other risk development protocols, multivariable logistic regression was performed to identify potential predictors of cardiovascular events.^40,41^ All independent variables were included in the model. Variables with a p-value greater than 0.10 were removed from the model unless they were statistically identified as confounders.

Confounders were determined as variables whose removal from the model led to a 20% or greater change in parameter estimates for one or more of the other variables, compared to the original model. The final model included confounders and all variables with a p-value of 0.10 or less.

### Risk Index Construction

The items to be included in the final index were chosen based on the statistical significance of their association with cardiovascular events in the logistic regression model, to strengthen the usability of the risk score and the practical need for a concise instrument that can be easily administered by healthcare professionals. To balance the scientific and statistical considerations of each variable, the following were considered during the risk score item selection process including the strength of association, confirmation of the variable as a risk factor in published literature, generalizability to the population, and the feasibility of obtaining valid and reliable information for each item in the risk score.

Point values for each of the risk questionnaire items were calculated by multiplying the β coefficients generated from regression analysis by 10 and rounding to the nearest integer. The calculated risk index scores were used in a multiple logistic regression model to predict probabilities of experiencing the outcome of a cardiovascular event.

Receiver operating curves (ROC) and corresponding C-statistics were utilized to determine the model’s discrimination ability between individuals with and without the outcome of interest.^42^

To validate the risk index, the generated predicted probabilities were separated into deciles. Ten risk classes were created based on the observed occurrence of cardiovascular events. The number of patients, the average predicted probability of the outcome, and observed incidence of events were calculated for each risk class. All statistical analyses were performed using R. This study followed the Strengthening the Reporting of Observational Studies in Epidemiology (STROBE) reporting guideline.^43^ This study was determined to be exempt by the Defense Health Agency Institutional Review Board.

## Results

### Descriptive Statistics

Baseline characteristics, including demographics (age, race, and sex), predefined risk factors (comorbidities), prescription drug information (including NSAID-related factors), and time since NSAID exposure are shown in Table 1. As described in Forbes et al, unadjusted analyses showed that cases of nonsteroidal anti-inflammatory drug-induced cardiovascular events (NAÏVE) were more likely to be older, non-white or black, male sex, and have a greater burden of illness as indicated by the number of comorbidities.^33^ Additionally, cases had higher numbers of previous cardiovascular events (history of myocardial infarction and cerebrovascular accidents). Cases were more frequently prescribed other potentially relevant medications including aspirin and anticoagulants than controls. Lastly, low/medium dose NSAIDs were prescribed more in both cases and controls compared to high-dose NSAIDs.

**Table 1.** Baseline Demographics & Predefined Risk Factors.

| Characteristics | Cases<br>(n = 231,967)<br>n (%) |  | Controls<br>(n = 2,319,670)<br>n (%) |  | P Value |
| --- | --- | --- | --- | --- | --- |
| **DEMOGRAPHICS** |  |  |  |  |  |
| Age (years), median (IQR) | 69 | 14 | 49 | 32 | <0.001 |
| Age Group (years) |  |  |  |  | <0.001 |
| 18-34 | 3971 | 1.71 | 673864 | 29 |  |
| 35-44 | 6444 | 2.78 | 346182 | 14.9 |  |
| 45-54 | 18485 | 7.97 | 334967 | 14.4 |  |
| 55-64 | 54305 | 23.4 | 404898 | 17.5 |  |
| 65-74 | 89380 | 38.5 | 414475 | 17.9 |  |
| 75+ | 59382 | 25.6 | 145284 | 6.26 |  |
| Male | 216417 | 93.3 | 1916362 | 82.6 | <0.001 |
| Race |  |  |  |  | <0.001 |
| Black or African American | 47039 | 20.3 | 5000047 | 21.6 |  |
| White | 145523 | 62.7 | 1553051 | 67 |  |
| Other | 39405 | 17 | 266572 | 11.5 |  |
| **PREDEFINED RISK FACTORS** |  |  |  |  |  |
| Diabetes | 96042 | 41.4 | 359266 | 15.5 | <0.001 |
| Hypertension | 181387 | 78.2 | 829599 | 35.8 | <0.001 |
| Dyslipidemia | 148788 | 64.1 | 773176 | 33.3 | <0.001 |
| History of Myocardial Infarction | 13314 | 5.74 | 14497 | 0.625 | <0.001 |
| Arthritis or Spondylitis | 6887 | 2.96 | 31240 | 1.35 | <0.001 |
| Peripheral Arterial Disease | 21595 | 9.31 | 38968 | 1.68 | <0.001 |
| Chronic Kidney Disease | 29643 | 12.8 | 64783 | 2.79 | <0.001 |
| Atherothrombotic Disease | 10885 | 4.69 | 20250 | 0.873 | <0.001 |
| History of Tobacco Use (Dx Only) | 34515 | 14.9 | 190430 | 8.21 | <0.001 |
| Cerebrovascular Disease | 41791 | 18 | 38342 | 1.65 | <0.001 |
| Coronary Artery Disease | 69186 | 29.8 | 126074 | 5.43 | <0.001 |
| Cardiomyopathy | 16127 | 6.95 | 17439 | 0.752 | <0.001 |
| Aspirin Use | 69054 | 29.8 | 170292 | 7.34 | <0.001 |
| Other Anticoagulant Use | 38386 | 16.5 | 52462 | 2.26 | <0.001 |
| Obstructive Sleep Apnea | 56751 | 24.5 | 357819 | 15.4 | <0.001 |
| Liver Dysfunction | 7497 | 3.23 | 26553 | 1.14 | <0.001 |
| COPD | 50860 | 21.9 | 119186 | 5.14 | <0.001 |
| **PRESCRIPTION DRUG INFORMATION** |  |  |  |  |  |
| BY ACTIVE INGREDIENT |  |  |  |  |  |
| Celecoxib | 17587 | 7.58 | 106505 | 4.59 | <0.001 |
| Diclofenac | 19681 | 8.48 | 124792 | 5.38 | <0.001 |
| Diflunisal | 101 | 0.0435 | 369 | 0.0159 | <0.001 |
| Etodolac | 6841 | 2.95 | 39324 | 1.7 | <0.001 |
| Fenoprofen | 80 | 0.0345 | 349 | 0.015 | <0.001 |
| Flurbiprofen | 60 | 0.0259 | 230 | 0.00992 | <0.001 |
| Ibuprofen | 69068 | 29.8 | 815442 | 35.2 | <0.001 |
| Indomethacin | 10302 | 4.44 | 49567 | 2.14 | <0.001 |
| Ketoprofen | 37 | 0.016 | 181 | 0.0078 | <0.001 |
| Ketorolac | 198 | 0.0854 | 1181 | 0.0509 | <0.001 |
| Mefenamic Acid | 3 | 0.0013 | 88 | 0.00379 | 0.08179 |
| Meloxicam | 62405 | 26.9 | 392807 | 16.9 | <0.001 |
| Nabumetone | 1543 | 0.665 | 7784 | 0.336 | <0.001 |
| Naproxen | 53190 | 22.9 | 496578 | 21.4 | <0.001 |
| Piroxicam | 526 | 0.227 | 4170 | 0.18 | <0.001 |
| Sulindac | 2451 | 1.06 | 11862 | 0.511 | <0.001 |
| Other (Summation of Highlighted) | 2540 | 1.09 | 14312 | 0.617 | <0.001 |
| BY SELECTIVITY |  |  |  |  |  |
| COX 1 | 82151 | 35.4 | 878473 | 37.9 | <0.001 |
| COX 2 | 86072 | 37.1 | 532371 | 23 | <0.001 |
| Non-Selective | 71888 | 31 | 614930 | 26.5 | <0.001 |
| BY DOSE CATEGORY |  |  |  |  |  |
| Low/Medium Dose | 189875 | 81.9 | 1656424 | 71.4 | <0.001 |
| High Dose | 44190 | 19.1 | 300509 | 13 | <0.001 |
| TIME SINCE INITIAL EXPOSURE (DAYS) |  |  |  |  | <0.001 |
| LEQ 30 | 15630 | 6.74 | 136896 | 5.9 |  |
| 31-90 | 33182 | 14.3 | 260316 | 11.2 |  |
| 91-180 | 38287 | 16.5 | 319594 | 13.8 |  |
| 181-365 | 52383 | 22.6 | 466322 | 20.1 |  |
| 366-730 | 54133 | 23.3 | 459522 | 19.8 |  |
| 731+ | 32563 | 14 | 257109 | 11.1 |  |
| No Exposure | 5789 | 2.5 | 419911 | 18.1 |  |

**Table 2.**
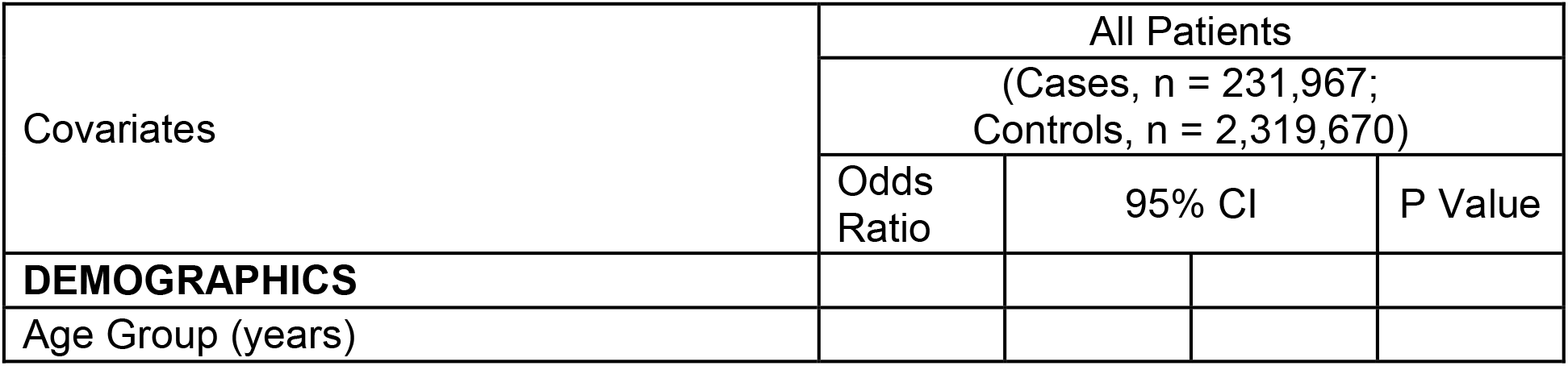
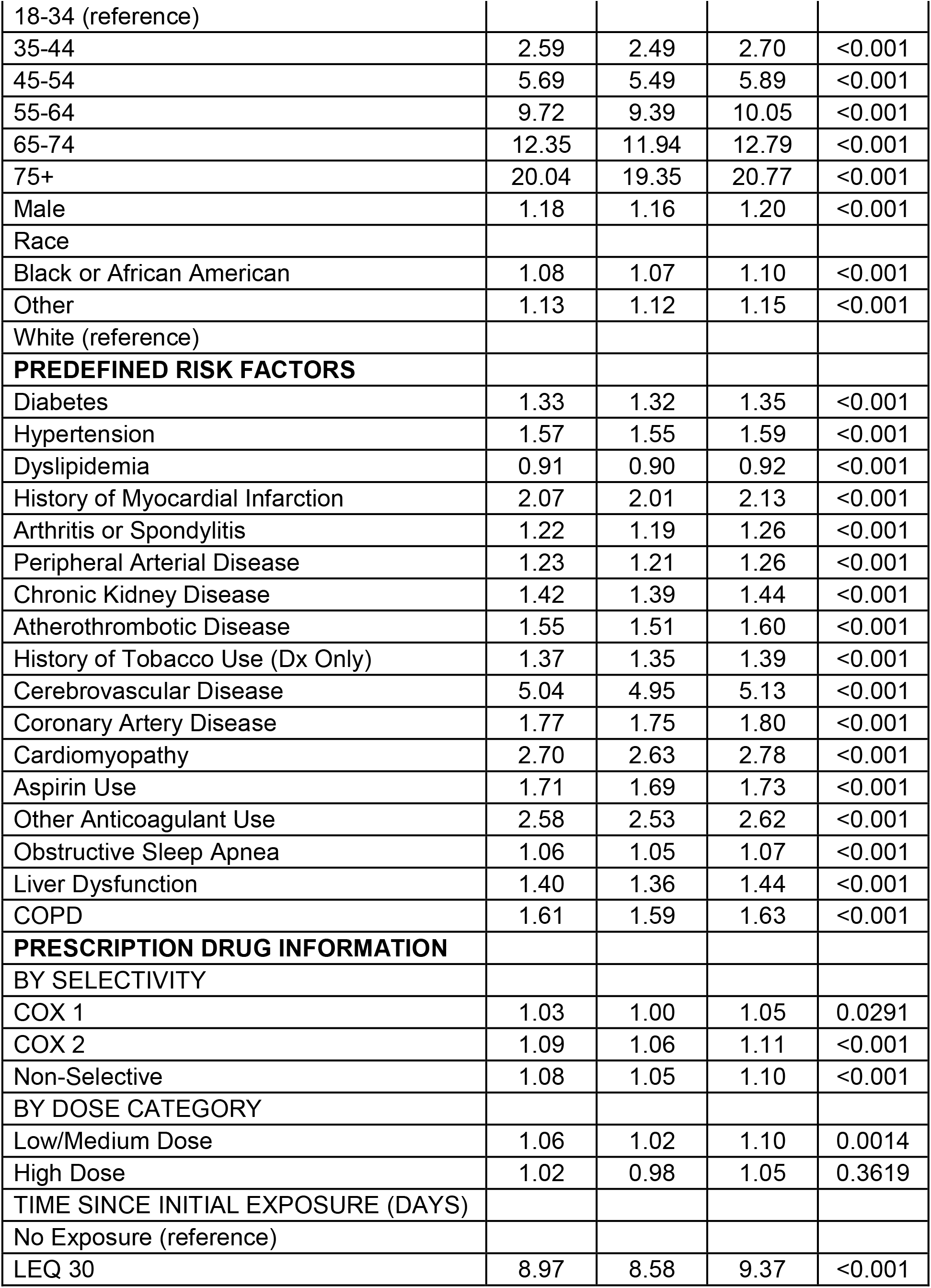
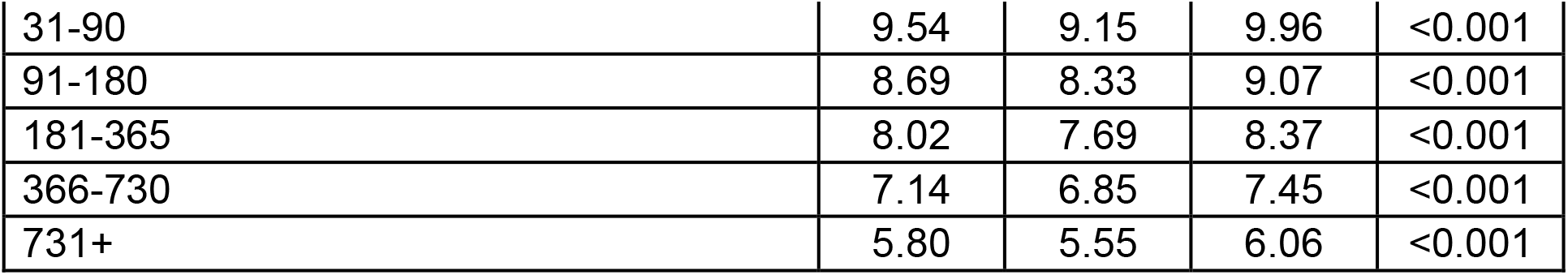
Primary Outcome: Composite Cardiovascular Events.

### Multivariable Modeling

The logistic regression model for the primary outcome of NSAID-related composite cardiovascular event of myocardial infarction, nonfatal cerebrovascular accident, and new heart failure resulted in multiple, independent, statistically significant associations.^33^ Of note, “History of heart failure” was removed from the predefined risk factors as there were only 10 cases of prior heart failure despite the large sample size.

High-dose NSAID use was the only independent variable excluded from multivariable regression modeling due to P>0.10 on bivariate analysis. Demographic variables associated with higher odds of nonsteroidal anti-inflammatory drug-induced cardiovascular events (NAÏVE) included age 45-54 (OR 5.69, 95% CI 5.49, 5.89), 55-64 (OR 9.72, 95% CI 9.39, 10.05), 65-74 (OR 12.35, 95% CI 11.94, 12.79), and 75 and older (OR 20.04, 95% CI 19.35, 20.77). Multiple predefined risk factors were associated with an event including: cerebrovascular disease (OR 5.04, CI 95% 4.95, 5.13), cardiomyopathy (OR 2.70, 95% CI 2.63, 2.78), history of myocardial infarction (OR 2.07, 95% CI 2.01, 2.13), coronary artery disease (OR 1.77, 95% CI 1.75, 1.80), COPD (OR 1.61, 95% CI 1.59, 1.63), and hypertension (OR 1.57, 95% CI 1.55, 1.59).

NSAID exposure was the NSAID-specific risk factor with the strongest association to NAÏVE and the highest likelihood peaking after 31-90 days of exposure (OR 9.54, 95% CI 9.15, 9.96), followed by less than 30 days (OR 8.97, 95% CI 8.58, 9.37), and 91-180 days (OR 8.69, 95% CI 8.33, 9.07) with a continued trend of slowly declining risk with longer term exposure. Medication-related risk factors associated with higher odds of the outcome were aspirin (OR 1.71, 95% CI 1.69, 1.73) and other anticoagulants (OR 2.58, 95% CI 2.53, 2.62).^33^ The odds ratios of secondary outcomes are summarized in Appendix Table 1.

### NAÏVE Risk Index Score

Table 3 shows the risk index items retained from the statistically significant predictors in the final model, and their corresponding assigned point values. Several statistically significant items in the final model were excluded in the final risk score due to lack of clinical significance, and overall impact on model simplicity and accuracy.

**Table 3.** Risk Score Calculation.

| Question | Points for Yes Response |
| --- | --- |
| **Patient's age group** |  |
| 35 – 44 | 9 |
| 45 – 54 | 17 |
| 55 – 64 | 23 |
| 65 – 74 | 25 |
| 75+ | 30 |
| **In the past year, has the patient had a healthcare visit (outpatient, inpatient, or ED) involving any of the following health conditions?** |  |
| Diabetes | 3 |
| Hypertension | 5 |
| Myocardial infarction | 7 |
| Chronic kidney disease | 3 |
| Atherothrombotic disease | 4 |
| History of tobacco use | 3 |
| Cerebrovascular disease | 16 |
| Coronary artery disease | 6 |
| Cardiomyopathy | 10 |
| Liver dysfunction | 3 |
| COPD | 5 |
| **Other medications** |  |
| Aspirin | 5 |
| Other anticoagulant | 9 |
| **Time since initial NSAID exposure (Prescription $\geq 7$ days and no gaps $> 30$ days)** |  |
| 30 days or fewer | 22 |
| 31 – 90 days | 23 |
| 91 days or more | 21 |

Demographics excluded from the final risk score were sex and race. Predefined risk factors (comorbidities) excluded were history of heart failure, obstructive sleep apnea (OSA), dyslipidemia, peripheral arterial disease, and arthritis/spondylosis. Excluded NSAID specific factors were COX selectivity and dose.

Table 4 presents risk classes by deciles of predicted probability of NAÏVE and the corresponding observed incidence. Based on risk factors present/absent during the past 12 months before the index date of the NAÏVE event, the predicted probability of a cardiovascular event ranged from 3% in the lowest risk class to 93% in the highest, and the observed incidence of NAIVE increased commensurately. The risk class model’s C-statistic was 0.88 and Hosmer-Lemeshow goodness-of-fit statistic 1723 (P<0.001), indicating very good calibration and discrimination between patients with and without an event (Table 4).

**Table 4.** Risk Classes and Predicted Probabilities of NSAID-Induced Cardiovascular Events.

| **Risk Class** | **Risk Index Score (Points)** | **All Patients (n=2551637), n(%)\*** | **Average Predicted Probability (95%CI)** | **Observed Incidence** |
| --- | --- | --- | --- | --- |
| 1 | 0-50 | 1,811,997 (71.0) | 0.03 (0.03, 0.03) | 0.02 |
| 2 | 51-58 | 409,312 (16.0) | 0.14 (0.14, 0.14) | 0.14 |
| 3 | 59-63 | 137,973 (5.4) | 0.24 (0.24, 0.24) | 0.26 |
| 4 | 64-68 | 77,499 (3.0) | 0.34 (0.34, 0.34) | 0.39 |
| 5 | 69-72 | 37,932 (1.5) | 0.46 (0.46, 0.46) | 0.49 |
| 6 | 73-76 | 27,188 (1.1) | 0.55 (0.55, 0.55) | 0.55 |
| 7 | 77-80 | 18,131 (0.7) | 0.65 (0.65, 0.65) | 0.60 |
| 8 | 81-85 | 15,088 (0.6) | 0.74 (0.74, 0.74) | 0.64 |
| 9 | 86-93 | 11,443 (0.4) | 0.84 (0.83, 0.84) | 0.69 |
| 10 | >=94 | 5,074 (0.2) | 0.93 (0.93, 0.93) | 0.75 |

\*Percentage total not equal to 100 due to rounding
Model Performance
C-Statistic=0.88

## Discussion

NSAIDs have few alternatives and will continue to be utilized due to the strong body of evidence to support their use for a variety of inflammatory and arthritic conditions including osteoarthritis and low back pain which are among the most common chronic pain conditions where they remain first-line treatment.^20-26^ Healthcare providers are aware of the potentially significant adverse effects with NSAID medications and have strategies around mitigating risks for GI bleeds and monitoring for NSAID-related renal impairment, but not for cardiovascular risk due to an inability to characterize that risk and predict the probability in their patients.^9^ Frontline healthcare providers should have clarity around the level of risk for each of their patients given the widespread nature of NSAID use and the high incidence of cardiovascular events.

A novel screening tool was developed to estimate the risk of nonsteroidal anti-inflammatory drug-induced cardiovascular events (myocardial infarction, non-fatal stroke, and new heart failure) for patients prescribed oral NSAIDs. The NAÏVE risk scoring tool performed well in the VHA/DoD study sample in identifying patients at increased risk of such events. Higher risk scores correlated closely with increased observed occurrence of events. NAÏVE is the first instrument intended to provide healthcare professionals with clinical decision support for assessing the potential for the most serious of adverse effects that can occur in patients being treated for common pain and inflammatory conditions using NSAIDs. It provides current, quantitative, evidence-based information about a patient’s level of risk of serious prescription NSAID-induced cardiovascular events. NAÏVE, which is based on a multivariable regression model, integrates independent risk factors and adjusts for confounding influences. As a result, NAÏVE can provide valuable decision support to health care professionals seeking to improve the safe and effective use of NSAIDs for pain management, particularly in complex patients who are biologically vulnerable to cardiovascular events.

### Intended Use and Interpretation of Results

The NAÏVE risk scoring tool requires responding to 13 items divided into 4 sections that include factors well documented in the literature as predictors of cardiovascular events. They comprise the risk factors most strongly associated with NAÏVE including age, NSAID exposure, predefined comorbidities, concomitant prescribed medications, and previous cardiovascular events. The NAÏVE risk scoring tool supports but does not replace the health care provider’s judgement in clinical decision-making and provides the basis for transparent conversations and shared decision-making regarding CV risk associated with NSAID use.

The intent of developing the NAÏVE screening tool is to assist health care professionals who are considering prescribing NSAIDs, to assess a patient’s baseline risk or re-evaluate current risk of non-steroidal anti-inflammatory drug-induced cardiovascular events (NAÏVE). Due to its reliance on readily available demographic, medical diagnosis codes, and prescription information in the electronic health record, the NAÏVE screening tool can be easily incorporated into automated clinical decision support tools, dashboards, and artificial intelligence driven health care initiatives for easier adoption and implementation to improve quality of care and efficiency for busy providers. It also can be employed periodically during ongoing treatment to reevaluate risk based on changes in a patient’s clinical condition or medication regimen. Explaining a NAÏVE score to a patient creates an opportunity to discuss the benefits and risks associated with the use of NSAIDs. For example, the provider may begin a discussion with, “Patients with risk scores similar to yours (e.g., 61 points) “were predicted to have X% chance (e.g., 26%) “of experiencing a cardiovascular event such as heart attack, stroke, or develop heart failure.” “This can occur quickly with the highest risk within the first 90 days of treatment. While NSAIDs are first-line treatment and may be effective for your condition, we should consider these potential risks and discuss alternatives.”

### Strengths and Limitations

NAÏVE was developed using extensive administrative health care data in the U.S. VHA/DoD population. Limitations inherent to observational studies using administrative data include: 1) medical coding errors/misclassification of comorbidities and previous cardiovascular events; 2) lack of data on patient adherence to prescribed medications or utilization of over-the-counter (OTC) medications including NSAIDs; 3) potentially relevant family history or demographics. In addition, while our VHA/DoD cohort had a significant number of younger patients, women were less represented than the general population and the study sample might not accurately reflect the broader U.S. population of users of prescription NSAIDs. As such, the predictive ability of NAÏVE, like other tools derived from observational data, is subject to residual confounding by currently unknown or excluded contributory factors.

The final risk index score does not include all known risk factors, such as family history of cardiovascular events. In addition, some variables associated with an event in the VHA/DoD sample were excluded because they were either not clinically significant or did not contribute in a meaningful way to the predicted probability of cardiovascular events and the priority to simplify the model.

### Implications for Future Research

While NAÏVE performed well in the VHA and DoD patient populations, it should be assessed and further validated in a separate population that is more representative of U.S. users of prescription NSAIDs. NAÏVE will also benefit from prospective reliability and validity testing across a broad spectrum of patients. NAÏVE can be formatted for electronic administration via Web or mobile platform to improve its real-world deployment by enabling automated risk scoring, and calculation of risk class.

## Conclusion

NAÏVE is the first-known published risk score to provide current, evidence-based information to health care providers regarding the risk of cardiovascular events with use of prescription NSAIDs. Its performance should be validated, and refined as necessary, in a more generalized patient population or prospectively. Once validated, this index will assist health care professionals in identifying patients who are at increased risk of serious NSAID-induced cardiovascular events and help with decision-making regarding interventions or alternative treatment options to mitigate risk.

## Data Availability

Data from VAVINCI database integrating electronic health records, administrative data, and clinical data from both the Veterans Health Administration (VHA) and the Department of Defense (DoD). This is a permissioned dataset that can't be shared outside the VA but the methodology to repeat or confirm the data is provided

## Notes

### Competing Interest Statement

The authors have declared no competing interest.

### Clinical Trial

Retrospective nested case-control study using administrative data

### Funding Statement

All authors are federal government employees and received no funding for this study.

### Author Declarations

The study was determined to be exempt by the Defense Health Agency Institutional Review Board

